# Impact of Antiretroviral Therapy on Anemia, Hepatotoxicity, and Immune Reconstitution Inflammatory Syndrome in HIV-Tuberculosis Co-Infected Patients

**DOI:** 10.1101/2024.09.24.24314274

**Authors:** Priyanka Gupta, Anil Kumar Tripathi, Shikha Gupta, Abhishek Gupta

## Abstract

**Introduction:** HIV continues to pose a significant global health challenge, causing gradual immune system decline and, if left untreated, progressing to AIDS. Although antiretroviral therapy (ART) has transformed the treatment landscape for HIV, it is frequently associated with adverse effects, including anemia, hepatotoxicity, and immune-related issues such as Immune Reconstitution Inflammatory Syndrome (IRIS), particularly in patients co-infected with tuberculosis (TB).

**Materials and methods:** In this prospective cohort study, 400 ART-naive, HIV-positive patients undergoing anti-tuberculosis therapy (ATT) at King George’s Medical University, Lucknow, India, were enrolled. The study evaluated the occurrence of opportunistic infections (OIs), ART regimen effectiveness, and key laboratory markers, including CD4 counts and hemoglobin levels.

**Results:** TB emerged as the most common OI, affecting 20.25% of the participants, with anemia present in 85% of the cohort. The prevalence of IRIS was notably higher in patients with a positive tuberculin skin test (20.6%) compared to those with a negative test (7.4%). Patients with IRIS exhibited significantly lower CD4 counts and hemoglobin levels compared to those without IRIS.

**Conclusion:** This study underscores the importance of integrated management approaches for HIV patients co-infected with TB, focusing on the complex interaction between immune status, ART effectiveness, and comorbidities like anemia and hepatotoxicity. Properly addressing these factors is crucial for improving patient outcomes in this high-risk population.

## Introduction

Human Immunodeficiency Virus (HIV) remains a major global health challenge, gradually weakening the immune system and, if untreated, leading to acquired immunodeficiency syndrome (AIDS) (1). Antiretroviral therapy (ART) has revolutionized HIV treatment, significantly advancing management in recent decades. However, patients still face side effects like anemia, hepatotoxicity, and fluctuations in CD4 levels, which can impact both health and ART effectiveness. In HIV-positive tuberculosis (TB) patients, Immune Reconstitution Inflammatory Syndrome (IRIS) complicates treatment, characterized by an enhanced immune response to TB antigens after starting ART. IRIS can manifest as either unmasking TB-IRIS, where subclinical TB becomes symptomatic, or paradoxical TB-IRIS, where existing TB worsens post-ART initiation (2).

Anemia is frequently made worse in individuals with IRIS and is a common comorbidity of TB and HIV infections (3). In HIV, anemia can have multiple causes, including long-term inflammation, iron, vitamin B12, folate, and dietary deficiencies, as well as direct effects of the virus on haematopoiesis. Multifactorial factors, including dietary inadequacies, ART-related toxicities, chronic inflammation, and bone marrow suppression, contribute to the pathogenesis of anemia in TB-IRIS (4). Anemia may deteriorate these individuals’ quality of life and clinical results.

Hepatotoxicity, often known as liver toxicity, is a serious issue in HIV patient care. ART treatments can damage the liver and worsen pre-existing liver disorders, especially when they involve medications that the liver metabolises (5). Hepatotoxicity has been linked to medications including isoniazid, rifampicin, and protease inhibitors. Because of the increased inflammatory response in the context of IRIS, there is an increased risk of liver injury (6). The treatment of individuals with co-infections with HIV and TB is greatly influenced by the selection of ART regimen. But in order to prevent side effects and drug-drug interactions, the interaction between various ART treatments and anti-TB therapies needs to be carefully considered.

For those living with HIV, the CD4 count is a vital indicator of immunological function. An increased risk of getting IRIS is linked to low baseline CD4 counts (7). IRIS is a manifestation of an inflammatory response against latent TB antigens that can arise from the restoration of immunological function after ART. For the purpose of anticipating and controlling IRIS, tracking the fluctuations of CD4 count is crucial. Furthermore, the assessment of the importance of CD4 count can be complicated by anemia and hepatotoxicity. Due to its effects on immune response and bone marrow function, anemia can lower CD4 count (8). By changing the metabolism and efficacy of antiretroviral medications, hepatotoxicity from ART can also have an indirect impact on CD4 count.

Recent research has focused on improving our understanding of the mechanisms and predictors of TB-IRIS in order to develop better management strategies. In an effort to lessen toxicity and improve patient outcomes, research on optimising ART regimens is also ongoing. To elucidate the relationship between ART regimens, anemia, hepatotoxicity, and CD4 count in patients with TB-related immune suppression. Our goal is to provide a comprehensive overview of the variables influencing the onset and treatment of disease due to T infection by integrating clinical data and recent research discoveries. To enhance treatment plans and results for individuals who co-infect HIV and T, it is imperative to understand these relationships.

## Material and methods

This prospective cohort study enrolled 400 HIV-positive patients from the ART Centre at King George’s Medical University (KGMU), Lucknow, India. The participants were selected based on the following inclusion criteria: (i) diagnosed with HIV infection, (ii) eligible for ART according to National AIDS Control Organization (NACO) guidelines, (iii) ART-naive, (iv) HIV-positive patients currently on anti-tuberculosis therapy (ATT), and (v) willing to participate in the study. Patients were excluded if they (1) did not consent to participate, or (2) were already receiving ART.

### Diagnosis of Active TB

Active TB diagnosis required either two positive sputum smears for acid-fast bacilli (AFB), Participants were categorized into two groups: (1) HIV patients with active TB on ATT, and (2) HIV patients without active TB evidence.

### Laboratory methods

Blood samples for CD4 cell count measurement were collected at baseline and again at the six-month follow-up. CD4 cell counts were analysed using standard three-color flow cytometry (FACScan; Becton Dickinson, San Jose, CA).

### Data collection

All participants were confirmed HIV-positive using the Enzyme linked immunosorbent assay (ELISA) method. A pre-designed structured questionnaire was used to gather socio-demographic data, clinical characteristics, and information on risk factors for IRIS. Detailed clinical histories and thorough examinations were conducted for all patients.

### Ethical approval

This study was approved by the Institutional ethical committee of King George’s Medical University, Lucknow, India (Ref. Code: - XXXV ECM/B-P3, No.694-R. Cell-09). Written informed consent for the participation in the study was obtained prior to enrolment from all the participants. Only participants who had consented were enrolled in the study and all information including their HIV status was kept confidential.

### Statistical analysis

Continuous variables such as age, Hb and CD_4_ cell count were summarized into means and medians as appropriate with t test, paired and unpaired as appropriate. The 95% confidence intervals were constructed around the estimates and the p-values used as a measure of statistical significance. A p-value of 0.05 or less was considered significant. Data analysis was done using SPSS 15.0 version statistical software.

## Results

### Pattern of Opportunistic Infection

Opportunistic infections (OIs) are prevalent among HIV patients due to their compromised immune systems. The predominant OIs observed in the patients were TB: 20.25%, Oral candidiasis: 16.5%, Cryptococcal meningitis: 2.5%, Diarrhoea: 1.5%, Scabies and sexually transmitted diseases (STDs): 1.25%, Carcinoma cervix, herpes zoster, non-Hodgkin’s lymphoma (NHL), and Progressive Multifocal Leukoencephalopathy (PML): 0.5% each, HIV neuropathy and molluscum contagiosum: 0.25% each. Over half of the patients (54.25%) did not present with any of these OIs at the time of the study (Table 1).

**Table 1:** Distribution of patients by OIs.

| **Name of OI non-TB** | **No. (n=400)** | **Percent (%)** |
| --- | --- | --- |
| Candidiasis | 66 | 16.5 |
| Carcinoma cervix | 2 | 0.5 |
| Cryptococcal meningitis | 10 | 2.5 |
| Diarrhoea | 6 | 1.5 |
| Herpes zoster infection | 2 | 0.5 |
| HIV Nephropathy | 1 | 0.25 |
| Molluscum contagiosum | 1 | 0.25 |
| non-Hodkins Lymphoma | 2 | 0.5 |
| PML | 2 | 0.5 |
| Scabies | 5 | 1.25 |
| STD | 5 | 1.25 |
| TB | 81 | 20.25 |
| None | 217 | 54.25 |

OI, opportunistic infection; TB, tuberculosis; HIV, human immunodeficiency virus; PML, progressive multifocal leukoencephalopathy; STD, sexually transmitted diseases

### ART Regimens

The distribution of patients by ART regimen is presented in the Figure 1. About one third were on STV+LMV+EFV (32.5%) and ZDV+LMV+NVP (32%). However, 23.3% were on STV+LMV+NPV regimen. Only 12.3% were on ZDV+LMV+EFV regimen.

**Figure 1:**
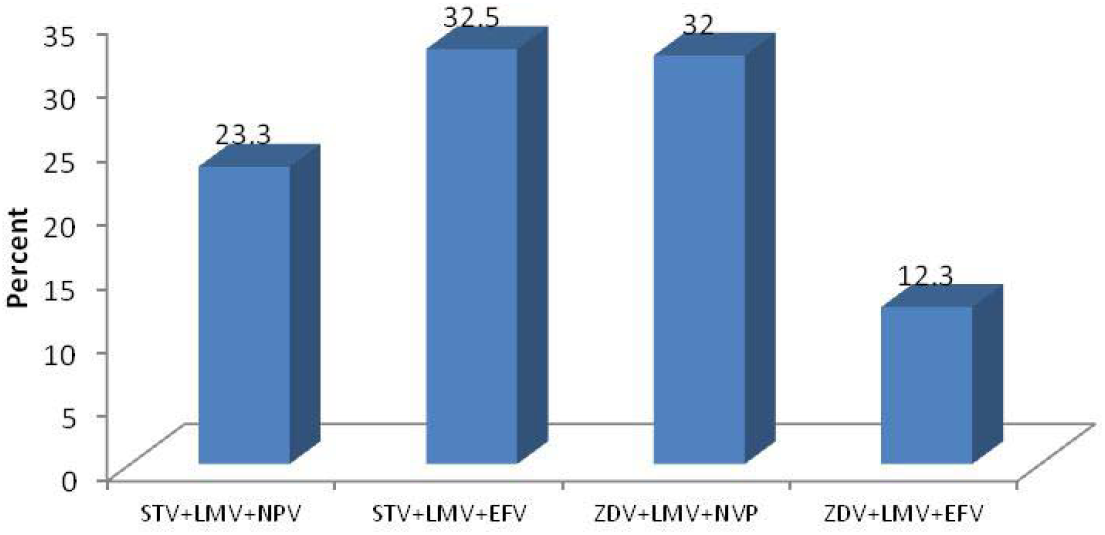
Distribution of patients by ART regimen.

### Prevalence of IRIS

The prevalence of IRIS was insignificantly lower in active TB patients (8.6%) than without TB patients (9.7%) (RR**=**0.89, 95% CI= 0.41-1.95, p=0.77). However, the prevalence was significantly higher among TST positive (20.6%) patients as compared to TST negative (7.4%) (RR=2.78, 95% CI= 1.51-5.14, p=0.001). hepatomegaly (23.3%, RR=2.79, 95% CI= 1.34-5.78, p=0.007) and Abdominal lymphadenopathy (35.3%, RR=4.22, 95% CI= 2.05-8.71, p<0.001) was also associated with higher risk of TB IRIS. There was no effect of VDRL and HBsAg and HCV results on the prevalence of IRIS (Table 2).

**Table 2:** Clinical Characteristics and Risk Factors.

| Variables | Number of patients<br>(n=400) | Prevalence of IRIS |  | RR (95% CI), p value |
| --- | --- | --- | --- | --- |
|  |  | No. | % |  |
| TB |  |  |  |  |
| Active TB | 81 | 7 | 8.6 | 0.89 (0.41-1.95), 0.77 |
| Without TB | 319 | 31 | 9.7 |  |
| TST |  |  |  |  |
| Positive | 63 | 13 | 20.6 | 2.78 (1.51-5.14), 0.001* |
| Negative | 337 | 25 | 7.4 |  |
| Hepatomegaly |  |  |  |  |
| Positive | 30 | 7 | 23.3 | 2.79 (1.34-5.78), 0.007* |
| Negative | 370 | 31 | 8.4 |  |
| Abdominal lymphadenopathy |  |  |  |  |
| Positive | 30 | 6 | 35.3 | 4.22 (2.05-8.71), <0.001* |
| Negative | 370 | 32 | 8.4 |  |
| VDRL |  |  |  |  |
| Positive | 32 | 3 | 9.4 | 0.99 (0.32-3.03), 0.98 |
| Negative | 368 | 35 | 9.5 |  |
| HBsAg |  |  |  |  |
| Positive | 10 | 1 | 10.0 | 1.05 (0.16-6.93), 0.96 |
| Negative | 390 | 37 | 9.5 |  |
| HCV |  |  |  |  |
| Positive | 8 | 1 | 12.5 | 1.32 (0.21-8.50), 0.77 |
| Negative | 392 | 37 | 9.4 |  |

IRIS, Immune reconstitution inflammatory syndrome; TB, tuberculosis; TST, tuberculin skin test; VDRL, venereal disease research laboratory; HBsAg, hepatitis B surface antigen; HCV, hepatitis C virus; RR, relative risk; CI, confidence interval.
\*Significant p value (<0.05)

**Table 3:**
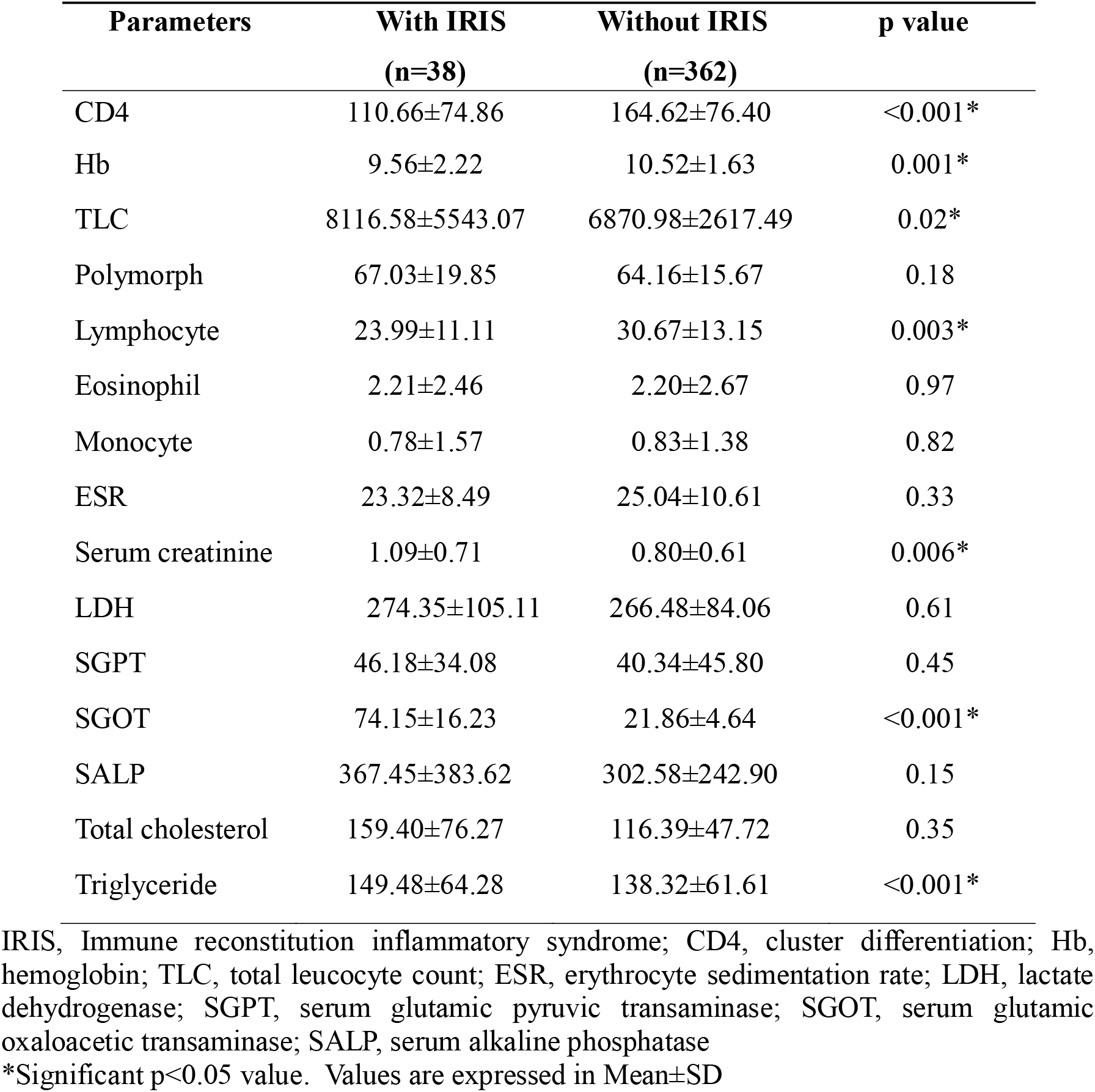
Comparison of baseline biochemical parameters between development of IRIS and Without IRIS.

### Biochemical parameters

A comparison of baseline biochemical parameters between patients with and without immune reconstitution inflammatory syndrome (IRIS) shows significant differences in several key metrics. CD4 count was notably lower in the IRIS group (110.66±74.86) compared to those without IRIS (164.62±76.40, p<0.001). Hemoglobin (Hb) levels were also lower in the IRIS group (9.56±2.22 vs. 10.52±1.63, p=0.001), while total leukocyte count (TLC) was higher (8116.58±5543.07 vs. 6870.98±2617.49, p=0.02). Lymphocyte percentages were lower in those with IRIS (23.99±11.11 vs. 30.67±13.15, p=0.003), and serum creatinine levels were elevated (1.09±0.71 vs. 0.80±0.61, p=0.006). Additionally, serum glutamic-oxaloacetic transaminase (SGOT) and triglyceride levels were significantly higher in the IRIS group (p<0.001 for both). However, some parameters, such as polymorph counts, eosinophils, and monocytes, showed no significant differences between the groups.

## Discussion

HIV, a condition marked by immune system compromise and increased vulnerability to opportunistic infections (OIs), remains a critical global health concern. While antiretroviral therapy (ART) has significantly improved the prognosis for HIV patients, complications such as hepatotoxicity, anemia, and the development of immune reconstitution inflammatory syndrome (IRIS) continue to present challenges. This discussion explores the relationship between OIs, ART regimens, and biochemical markers influencing patient outcomes.

In our study, tuberculosis (TB) emerged as the most prevalent OI, affecting 20.25% of the cohort. According to the World Health Organization (1), TB remains one of the leading causes of illness and death among people living with HIV, consistent with our findings and previous literature. Oral candidiasis was the second most common OI, affecting 16.5% of the cohort, aligning with earlier studies that underscore the high incidence of mucosal infections in HIV patients (9). Less common OIs, such as herpes zoster (0.5%) and cryptococcal meningitis (2.5%), followed epidemiological patterns reported in earlier research. Notably, more than half of the patients (54.25%) did not present with OIs at the time of investigation, indicating that ART may have effectively reduced the prevalence of these infections. This supports existing evidence that modern ART regimens enhance immune function and reduce the risk of OIs (10).

Anemia, a common condition in the general population and particularly among HIV patients attending ART Centres (11), was prevalent in our cohort, affecting 85% of the patients. This finding is in line with *Kaudha et al*. study (12), which associated anemia with low hemoglobin (Hb) levels, and decreased CD4 counts, a well-established marker of poor clinical outcomes in HIV patients. The correlation between anemia and advanced HIV stages further emphasizes its prognostic importance.

Anemia is also independently linked to higher mortality rates in HIV patients. Our study observed an association between anemia and an increased incidence of IRIS. Although the underlying mechanisms remain unclear, factors contributing to anemia, such as poor nutrition, low CD4 counts, high viral loads, and inflammatory cytokines, may impair erythropoietin production and red blood cell formation. These factors could potentially serve as predictors for a rapid immune recovery (i.e., IRIS) following ART initiation. Thus, early diagnosis and treatment of anemia may mitigate the risk of IRIS.

We examined the association between ART regimens and IRIS, focusing on patients receiving one of four combinations: two Nucleoside Reverse Transcriptase Inhibitors (NRTI) and one Non-Nucleoside Reverse Transcriptase Inhibitor (NNRTI). No specific ART regimen was found to be associated with a higher incidence of IRIS, corroborating previous studies (13,14). The distribution of ART regimens was as follows: 32.5% received Stavudine + Lamivudine + Efavirenz (STV+LMV+EFV), 32% received Zidovudine + Lamivudine + Nevirapine (ZDV+LMV+NVP), 23.3% received Stavudine + Lamivudine + Nevirapine (STV+LMV+NPV), and 12.3% received Zidovudine + Lamivudine + Efavirenz (ZDV+LMV+EFV). These regimens are commonly selected for their effectiveness and tolerability, reflecting the need for personalized treatment approaches (15).

Our findings indicated that patients with positive tuberculin skin test (TST) results had a higher incidence of IRIS (20.6%) compared to those with negative TST results (7.4%). This supports previous research showing that latent TB increases the risk of IRIS after ART initiation (13,16). Additionally, hepatomegaly and abdominal lymphadenopathy were associated with a higher risk of IRIS, consistent with earlier studies (17). However, no significant associations were found between IRIS and markers such as VDRL, HBsAg, and HCV, in line with recent research suggesting that these markers may not be critical in predicting IRIS.

During the one-year follow-up, no cases of IRIS-associated hepatitis were reported. However, a significant increase in serum transaminases (SGOT, SGPT) was observed at the onset of IRIS, consistent with previous findings (18). Although nevirapine is commonly associated with hepatotoxicity, our data did not point to a drug-induced cause, as similar increases were observed in patients on efavirenz. Elevated transaminases can impair liver function, indicating that caution is needed when initiating ART in patients with pre-existing liver conditions.

The rapid PCR method has become the preferred diagnostic tool for TB detection, and enhancing the development of rapid molecular diagnostics for subclinical HIV co-infections could aid in controlling the TB epidemic (19). Misdiagnosis of smear-negative TB based solely on clinical presentation can lead to inappropriate treatment, increasing the risk of adverse treatment reactions and drug resistance due to the overlapping clinical features of various OIs (20).

Effective ART administration and high adherence rates are critical in managing HIV and preventing TB, particularly in co-infected individuals (21). Lastly, our study identified a strong association between IRIS and elevated levels of serum creatinine, total cholesterol, and total lymphocyte count (TLC). These findings highlight the complex interactions between lipid metabolism, renal function, and systemic inflammation in HIV patients. Elevated cholesterol may signal metabolic abnormalities related to HIV or ART, while increased serum creatinine and TLC could indicate ongoing inflammation or renal impairment.

## Conclusion

This study highlights the complexities of managing HIV in patients co-infected with tuberculosis (TB). While antiretroviral therapy (ART) improves outcomes, challenges such as Immune Reconstitution Inflammatory Syndrome (IRIS), anemia, and hepatotoxicity persist. TB, the most common opportunistic infection, increases the risk of IRIS, particularly in patients with low baseline CD4 counts, positive tuberculin skin tests, hepatomegaly, and abdominal lymphadenopathy. Anemia and liver dysfunction further contribute to poorer outcomes. These findings emphasize the importance of monitoring ART regimens and patient-specific risk factors to enhance management of HIV and TB.

## Data Availability

All data produced in the present study are available upon reasonable request to the authors.

## Acknowledgements

The patients’ participation in the study is greatly appreciated by the authors. The contributing physicians and residents from KGMU, Lucknow’s Department of Clinical Hematology and Medical Oncology for their kind assistance.

## References

1. World Health Organization (WHO). Guideline: Anemia in HIV-infected adults and adolescents. WHO. 2021.

2. Lanzafame M, Vento S. Tuberculosis-immune reconstitution inflammatory syndrome. J Clin Tuberc Other Mycobact Dis. 2016 Mar 11; 3: 6–9. doi: 10.1016/j.jctube.2016.03.002.

3. Ara Jo-Pereira M, Sheikh V, Sereti I, Barreto-Duarte B, Arriaga MÍB, Tib Rcio R, Vinhaes CL, Pinto-de-Almeida M, Wang J, Rupert A, Roby G, Shaffer D, Ananworanich J, Phanuphak N, Sawe F, Andrade BB. Association between severe anaemia and inflammation, risk of IRIS and death in persons with HIV: A multinational cohort study. eBioMedicine. 2022 Nov; 85:104309. doi: 10.1016/j.ebiom.2022.104309. Epub 2022 Oct 22.

4. Kerkhoff AD, Meintjes G, Opie J, Vogt M, Jhilmeet N, Wood R, Lawn SD. Anaemia in patients with HIV-associated TB: relative contributions of anaemia of chronic disease and iron deficiency. Int J Tuberc Lung Dis. 2016 Feb; 20(2):193–201. doi: 10.5588/ijtld.15.0558.

5. Gebremicael G, Tola HH, Gebreegziaxier A, Kassa D. Incidence of Hepatotoxicity and Factors Associated During Highly Active Antiretroviral Therapy in People Living with HIV in Ethiopia: A Prospective Cohort Study. HIV AIDS (Auckl). 2021 Mar 25; 13: 329–336. doi: 10.2147/HIV.S283076.

6. Ramappa V, Aithal GP. Hepatotoxicity Related to Anti-tuberculosis Drugs: Mechanisms and Management. J Clin Exp Hepatol. 2013 Mar; 3(1): 37–49. doi: 10.1016/j.jceh.2012.12.001. Epub 2012 Dec 20.

7. Vignesh R, Balakrishnan P, Tan HY, Yong YK, Velu V, Larsson M, Shankar EM. Tuberculosis-Associated Immune Reconstitution Inflammatory Syndrome-An Extempore Game of Misfiring with Defense Arsenals. Pathogens. 2023 Jan 29; 12(2): 210. doi: 10.3390/pathogens12020210.

8. McMichael AJ, Borrow P, Tomaras GD, Goonetilleke N, Haynes BF. The immune response during acute HIV-1 infection: clues for vaccine development. Nat Rev Immunol. 2010 Jan; 10(1): 11–23. doi: 10.1038/nri2674. Epub 2009 Dec 11.

9. Taverne-Ghadwal L, Kuhns M, Buhl T, Schulze MH, Mbaitolum WJ, Kersch L, Weig M, Bader O, Groß U. Epidemiology and Prevalence of Oral Candidiasis in HIV Patients From Chad in the Post-HAART Era. Front Microbiol. 2022 Feb 17; 13: 844069. doi: 10.3389/fmicb.2022.844069.

10. Mofenson LM, Brady MT, Danner SP, Dominguez KL, Hazra R, Handelsman E, Havens P, Nesheim S, Read JS, Serchuck L, Van Dyke R; Centers for Disease Control and Prevention; National Institutes of Health; HIV Medicine Association of the Infectious Diseases Society of America; Pediatric Infectious Diseases Society; American Academy of Pediatrics. Guidelines for the Prevention and Treatment of Opportunistic Infections among HIV-exposed and HIV-infected children: recommendations from CDC, the National Institutes of Health, the HIV Medicine Association of the Infectious Diseases Society of America, the Pediatric Infectious Diseases Society, and the American Academy of Pediatrics. MMWR Recomm Rep. 2009 Sep 4; 58(RR-11): 1–166.

11. Cao G, Wang Y, Wu Y, Jing W, Liu J, Liu M. Prevalence of anemia among people living with HIV: A systematic review and meta-analysis. eClinicalMedicine. 2022 Jan 26; 44: 101283. doi: 10.1016/j.eclinm.2022.101283.

12. Kaudha R, Amanya R, Kakuru D, Muhumuza Atwooki R, Mutebi Muyoozi R, Wagubi R, Muwanguzi E, Okongo B. Anemia in HIV Patients Attending Highly Active Antiretroviral Therapy Clinic at Hoima Regional Referral Hospital: Prevalence, Morphological Classification, and Associated Factors. HIV AIDS (Auckl). 2023 Oct 12; 15: 621–632. doi: 10.2147/HIV.S425807.

13. Quinn CM, Poplin V, Kasibante J, Yuquimpo K, Gakuru J, Cresswell FV, Bahr NC. Tuberculosis IRIS: Pathogenesis, Presentation, and Management across the Spectrum of Disease. Life. 2020; 10: 262. 10.3390/life10110262.

14. Lai RP, Meintjes G, Wilkinson RJ. HIV-1 tuberculosis-associated immune reconstitution inflammatory syndrome. Semin Immunopathol. 2016 Mar; 38(2): 185–198. doi: 10.1007/s00281-015-0532-2. Epub 2015 Sep 30.

15. Sinha S, Gupta K, Tripathy S, Dhooria S, Ranjan S, Pandey RM. Nevirapine-versus Efavirenz-based antiretroviral therapy regimens in antiretroviral-naive patients with HIV and Tuberculosis infections in India: a multi-centre study. BMC Infect Dis. 2017 Dec 11; 17(1): 761. doi: 10.1186/s12879-017-2864-0.

16. Gupta P, Tripathi AK, Jain A, Singh KP, Misra RP. Redefining the role of Tuberculin skin test of TB-IRIS in HIV Positive patients on antiretroviral therapy in North Indian Population. Indian J Prev Soc Med. 2012; 43(2): 183–187.

17. Bosamiya SS. The immune reconstitution inflammatory syndrome. Indian J Dermatol. 2011 Sep-Oct; 56(5): 476–479. doi: 10.4103/0019-5154.87114.

18. Huruy K, Mulu A, Mengistu G, Shewa-Amare A, Akalu A, Kassu A, Andargie G, Elias D, Torben W. Immune reconstitution inflammatory syndrome among HIV/AIDS patients during highly active antiretroviral therapy in Addis Ababa, Ethiopia. Jpn J Infect Dis. 2008 May; 61(3): 205–209.

19. Gupta P, Singh KP, Tripathi AK, Jain A, Prasad R. Role of polymerase chain reaction as a diagnostic tool in pulmonary tuberculosis. J Rec Adv Appl Sci. 2013; 28:19–24.

20. Gupta P, Gupta A, Singh KP. Tuberculosis Diagnosis in Patients Co-Infected with HIV: A Review. Austin J HIV/AIDS Res. 2024; 10(1): 1057.

21. Gupta P, Tripathi AK, Jain A, Prasad R, Singh KP, Vaish AK, Misra RP. Effect of IRIS development on survival in HIV-TB patients on antiretroviral therapy among north Indian population. Indian J Community Health. 2012; 24(2): 129–133.

